# Network analysis of cross-income-level collaboration on non-communicable disease: the example of multiple myeloma in Sub-Saharan Africa

**DOI:** 10.1101/2024.10.24.24316045

**Authors:** Kaiyi Yang, Sam Benkwitz-Bedford, Jean-Baptiste Cazier, Fabian Spill

## Abstract

Cross-income-level collaboration (CILC) is crucial for developing global health approaches that benefit low- and middle-income countries (LMICs). Multiple myeloma (MM) is a representative example of a complex, understudied disease in sub-Saharan Africa (SSA). Based on publications, we developed a network analysis tool to assess scientific collaborations. Here, we present findings from a systematic analysis of publications retrieved from PubMed between January 2002 and June 2022. We evaluated individual institutional contributions and collaboration patterns using undirected weighted networks. Our findings reveal that intraincome-level collaborations dominate MM research in SSA, with high-income countries (HICs) primarily engaging with a few local institutions, mainly in South Africa and Nigeria. Increasing CILC is essential to advance research in this area. Our analysis tool provides insights into the collaboration strength, highlights gaps in the field and identifies leading institutions, ultimately aiming to support the development of more effective international collaboration and research strategies in MM.

## 1 Background

Medical interventions are largely developed in high-income countries (HICs), and validated by clinical trials with cohorts whose population is rarely representative of the global population[1][2]. Research demonstrates that for many complex diseases such as cancer, patient characteristics including ethnicity strongly affect drug efficacy. Advancing global health, especially the health of poor and vulnerable populations across diverse geographic regions particularly in low-and-middle-income countries (LMICs) remains challenging[3][4]. The World Health Organization also reports that there remain persistent and widening gaps between those with the best and worst health and well-being[5]: poorer populations systematically experience worse health than richer ones. For example, there is a difference of 18 years of life expectancy between HICs and LMICs[5]. Each year, 17 million people die from non-communicable diseases (NCDs) before the age of 70; 86% of these premature deaths occur in LMICs. Of all NCD deaths, 77% are in LMICs. Cancers account for the second most common cause of NCD deaths.[6]

Sub-Saharan Africa is experiencing a marked increase in cancer burden, with more than 1 million incidents of cancers and nearly 800,000 cancer-related deaths projected in the year 2030, representing an approximately 85% increase from 2008[7]. Of all cancers occurring in SSA, hematologic malignancies have emerged as one of the major causes of morbidity and mortality[7].

Multiple myeloma (MM) is one of the most common hematological malignancies in SSA LMICs[8]. The mortality due to MM in the region is expected to increase by 90% from 2008 until 2030. This is the highest increase of the four major haematological diseases (Non-Hodgkin lymphoma, leukemia, Hodgkin lymphoma and MM)[9][10]. Incidence of MM is highly variable among countries but has increased uniformly since 1990, with the largest increase in middle and low-middle socio-demographic index countries[11].

Despite some heterogeneity, the prevalence of MM is challenging on the African continent, where, for example, it accounts for about 8.2% of all haematological malignancies in the Niger-Delta area [12][13].

The World Health Organization also points out that collaborations across sectors of healthcare and disciplines are key to the development of global health programs and that ultimately they are likely to lead to faster and more sustainable interventions in LMICs[14]. Collaborations between HICs and LMICs (cross-income-level collaboration) can involve exchanging skills and sharing expertise.[15] HICs are generally well-resourced and can contribute professional capabilities and specialised resources whilst LMIC partners contribute local clinical and other contextual knowledge.[16][17]Institutions and industries in the biomedical and pharmaceutical fields in HICs may also need information on potential partner institutions to help them conduct research and development of interventions that will impact African patients. This is necessary as drug effectiveness and toxicity can vary among ethnic groups.[18]. Collaborations with relevant LMICs will ensure that academic institutions and companies in HICs have an opportunity to assure that their interventions are inclusive and enhance global health[19]. More importantly, stronger research leadership in LMICs will help ensure that a country’s research agenda is aligned with its knowledge needs, and not merely with the research interests of HICs scientists[4]. It is therefore essential to identify the key institutions in the field, especially locally, and enhance their role through higher visibility worldwide.

In this work, we develop methods to infer collaboration networks and identify leading institutions focused on diseases in SSA, using MM as a representative example. We begin with a descriptive analysis at the country and institution levels, examining indicators to highlight key contributors. Network graphs are then visualized and analyzed at both levels. Finally, adjusted degree centrality algorithms are applied to identify top institutions in CILC, stratified by geographic location.

## 2 Methods

### 2.1 Data preparation

To identify leading countries and institutions working on MM in SSA, we focused on research papers containing affiliation information from the last 20 years (2002-2022). The list of sub-Saharan countries was obtained from a public website[20]. The search terms were ‘multiple myeloma’ AND ‘country’ (48 countries were listed in SSA) and ‘multiple myeloma’ AND ‘sub-Saharan’. After setting the database parameter to “pubmed” and searching for the keywords using the E-utilities (which are the public API to the NCBI Entrez system and allow access to all Entrez databases including PubMed, PMC, Gene, Nuccore and Protein[21][22]), we obtained 284 relevant publications from 1963 to 2022. Publications are not limited to English but their title, abstract, MeSH (Medical Subject Headings) or other fields should contain our search term in English. Publication types include reviews, observation studies, clinical trials, case reports etc. The lxml library in Python was then used to parse the publication website page and extract the relevant information (Website, PMID, affiliation, publication date, title, and list of authors). Since older publications do not provide information on affiliation, we included 154 publications containing precise affiliations between 2002 and 2022 for downstream analysis.

We used Python’s Geotext package to identify countries and cities from the ‘affiliation’ field. Missing information was often inferred and manually curated. For example, US institutions often omitted the country but listed the state or city, so we implemented a dictionary to match US states and cities to the country. We also used keyword searches (e.g., university, hospital) to extract institution names from long strings. Given that some institutions are under multiple names, we standardized the institution names using ROR (The Research Organization Registry) REST API[23][24], and for those that could not be identified correctly we used text filtering in Excel, approximate string matching ([25]) and manual checking. We manually curated part of 30% of the data. Finally, 65 countries and 408 institutions were identified.

The definitions of HIC in this article are taken from the World Bank’s website (according to the gross national income of each capital)[26]. All the countries that are not classified as HICs are LMICs i.e. there are only two levels in our dataset, HIC and LMIC.

### 2.2 Weighted network generation and visualization

The graphs G=(V, E) are defined by the sets of all vertices V and all edges E and are undirected weighted graphs[27] since no direction of collaboration is evident from the publication data.

We constructed two levels of network graphs: one at the country level and another at the institution level. Vertices represent countries or institutions, and edges indicate collaborative publications between them. If multiple authors from the same country or institution appear in a publication, a self-loop is generated.

Weights can be represented as a function *ω*: *E* → *R* that assigns each edge *e* ∈ *E* a weight *ω*_*E*_(*e*) or *ω*: *V* → *R* that assigns each vertex *v* ∈ *V* a weight *ω*_*V*_ (*v*).[28] The edge weights describe the strength of collaboration[29] while the vertex weights describe how active the country or institution is in the field. The exact weighting algorithm is as follows:

Each publication may list multiple affiliations, with different departments of the same institution resulting in different strings. For example, ‘School of Mathematics, University of Birmingham, UK’ and ‘Birmingham Medical School, University of Birmingham, UK’ are both considered as the University of Birmingham in the dataset.

If there are *N*_*P*_ publications, for the *k*^*th*^ publication, assume there are *n*_*k*_ affiliations (Table 1):

**Table 1:**
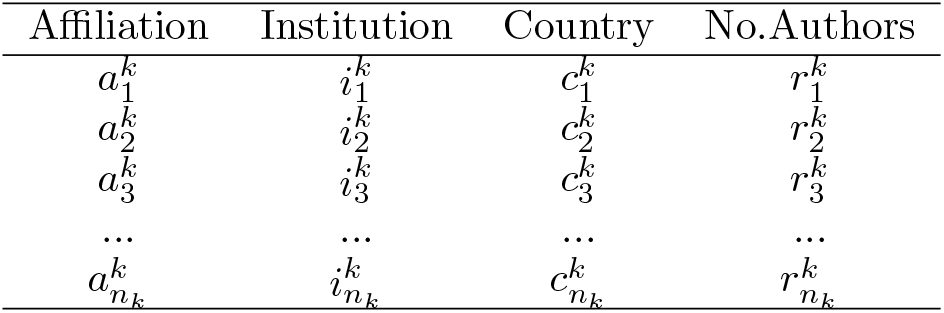
Affiliations of the kth publication.

| Affiliation | Institution | Country | No.Authors |
| --- | --- | --- | --- |
| $a_1^k$ | $i_1^k$ | $c_1^k$ | $r_1^k$ |
| $a_2^k$ | $i_2^k$ | $c_2^k$ | $r_2^k$ |
| $a_3^k$ | $i_3^k$ | $c_3^k$ | $r_3^k$ |
| ... | ... | ... | ... |
| $a_{n_k}^k$ | $i_{n_k}^k$ | $c_{n_k}^k$ | $r_{n_k}^k$ |

By combining the same institutions and summing the number of authors, we obtained a table at the institution level. Assume the number of institutions for the *k*^*th*^ publication is 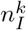, and the number of authors from the *j*^*th*^ institution 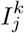 is 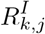.

By combining the same countries and summing the number of authors, we obtained a table at the country level. Assume the number of countries for the *k*^*th*^ publication is 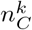, and the number of authors from the *q*^*th*^ country 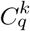 is 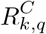.

For convenience, we introduced a level indicator *X. X* = *I* for institution level and *X* = *C* for country level and a matching function *δ* for strings *Str*_1_ and *Str*_2_ which represent the names of two institutions or countries:

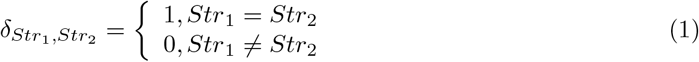

The weight (or size) of a vertex is the sum of the number of times the authors from the institution or country represented by that vertex have contributed to papers. Thus, if an author contributes to more than one paper, his contributions are counted multiple times. Assume the vertex is *X*_*m*_, and the weight of this vertex is *ω*_*V*_ (*X*_*m*_):

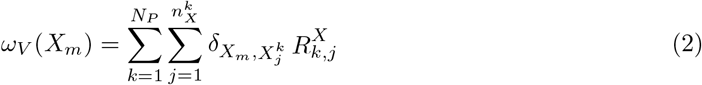

The strength of collaboration between the two different institutions or countries 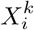 and 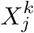 in the *k*^*th*^ publication 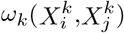 is increasing with the number of authors from either institution. For simplicity, we assume that the strength is given by the number of pairs of authors between each institution, which is the product of the number of authors 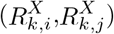,

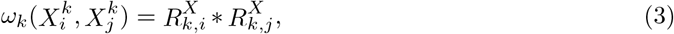

For authors from the same institution or country 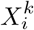, self-collaboration occurs if there is more than one author:

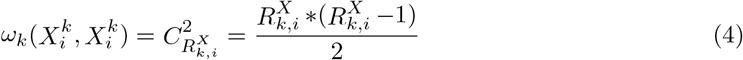

where *C* is the combination formula

After calculating the weights of collaboration between the institutions or countries in each publication, we obtain the weights of the edges between institutions or countries *X*_*m*_ and *X*_*n*_ is:

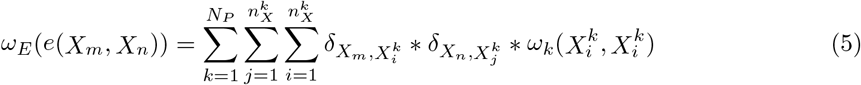

At the institution level, edges are classified into three categories based on the income levels of the institutions: LMLM (both in LMICs), LMHC (one in HIC and one in LMIC), and HCHC (both in HICs). This classification allows for calculating degrees for each category, reflecting the extent of an institution’s cooperation with different income-level institutions. It also aids in focusing on LMIC institutions and applying the adjusted degree centrality algorithm.

As we focus more on collaboration between institutions in HICs and those in LMICs, i.e. CILC, we have also analyzed bipartite graphs in the network graph section. A bipartite graph is a graph whose vertices can be divided into two disjoint and independent sets *L* and *H*, that is every edge connects a vertex in *L* to one in *H*. Vertex sets *L* and *H* are usually called the parts of the graph.[29] Here *L* is the set of LMICs and *H* is the set of HICs, so the bipartite graph only contains LMHC edges, and omits the self-loop and edges of cooperation between the same-income countries.

We refined and constructed an interactive tool to visualize the network. The tool is based on Python, pyvis and Jaal and can be found on interactive tool on Gitlab.

### 2.3 Intra- and inter-group cooperation: E-I index

We refer to collaborative ties between institutions in different income levels—in this case, between institutions in LMICs and institutions in HICs as external ties. Internal ties are collaborative ties between institutions in the same income-level countries. A classical measure of the pre-eminence of external over internal ties (or heterophily) is the E-I index by Krackhardt and Stern[30], which is given by

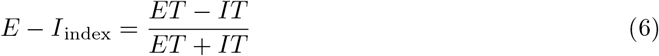

where ET and IT denote the total number of external and internal ties, respectively, in the context of this study, the E-I index is a measure of CILC. The index can be calculated either for the whole network, each group or each institution[31].

The E-I index ranges from -1 to 1. A value of -1 means all collaborations are internal (within LMICs or HICs), while 1 indicates all ties are cross-income-level (between LMICs and HICs). An index of 0 signifies an equal division of collaborations between the two groups.

For networks with nodes belonging to different groups, the researchers defined a regionalization index[32] and the nationalization index[33] based on the E-I index for different research purposes. The indexes above all consider only the number of internal and external ties. Given that our network is a weighted network, we have also defined the ‘strength regionalization index’ (SRI) that considers the strength of links based on the form of the E-I index:

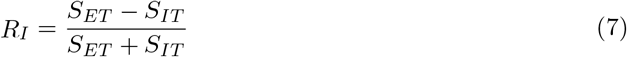

where *S*_*ET*_ is the sum of the strength of the external ties and *S*_*IT*_ is the sum of the strength of the internal ties. We applied this index to network data at the institutional level to examine the propensity of LMIC institutions to collaborate with HIC institutions, and vice versa.

### 2.4 Identifying leading institutions: betweenness centrality, eigenvector centrality, degree centrality and adjusted degree centrality at institution level

The ‘betweenness centrality’ [34] is based on the extent to which a particular node lies between other pairs of nodes in a network, connecting them. Therefore, nodes with high betweenness centrality are usually considered hubs in the network. The ‘eigenvector centrality’ [35] is based on the idea that a node is important if its neighbours are important.

Considering the specificity of the weighted collaborative networks we define, and our goal of identifying leading institutions in CILC, we define a few metrics and extend the degree centrality algorithm.

The total collaborative strength of a vertex (institution) *v*_*i*_, denoted by *S*(*v*_i_), is defined as the sum of the weights of all edges incident to it (including self-loop)[36], we denote *N* (*v*_*i*_) as the set of the neighbours of vertex *v*_*i*_, thus:

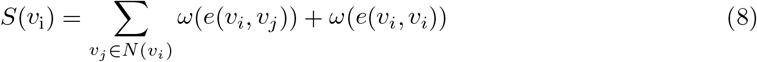

The second term in the equation 8 is equal to zero if *v*_*i*_ does not have a self-loop. The degree centrality of *v*_i_ ∈ *V* of an edge-weighted graph (*G, W*), denoted by 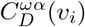 is defined by

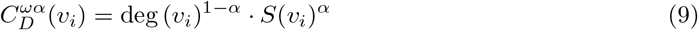

where *α* ∈ [0, 1]

The parameter *α* is called the tuning parameter. It is clear that when *α* = 0 then 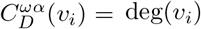 and when *α* = 1 then 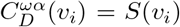). From the form of the function, it is easy to see that as the value of *α* increases, the degree centrality algorithm focuses more on the strength of the cooperation and ignores the degree.[37]

Firstly, we decompose the degree *deg*(*v*_*i*_) of each vertex (institution) *v*_*i*_ into two parts, the degree of the edges of the LMHC category denoted as deg_*LH*_ (*v*_*i*_), and the other degrees denoted as deg (*v*_*i*_) (including the degree of the HCHC category and the degree of the edges of class the LMLM category). So:

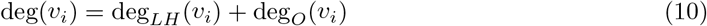

Secondly, the corresponding cooperative strengths *S*(*v*_i_) are also divided into two parts, the sum of the weights of the edges of the LMHC category noted as *S*_*LH*_ (*v*_*i*_), and the sum of the weights of the edges of the other categories noted as *S*_*O*_(*v*_*i*_) (including the sum of the weights of the edges of the HCHC category and the sum of the weights of the edges of the LMLM category). So:

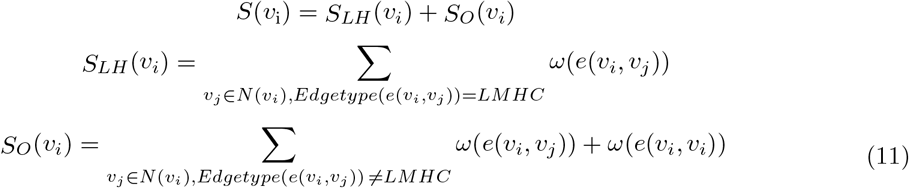

Given our previous classification of nodes and edges, we defined the split form of degree centrality:

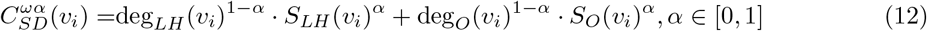

It maintains the property that when *α* = 0 then 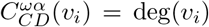 and when *α* = 1 then 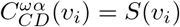.

Thirdly, without changing the properties of the above equation, we adjust the coefficients so that institutions with more cross-income-level collaborations have higher centrality scores. The adjusted degree centrality function is defined by:

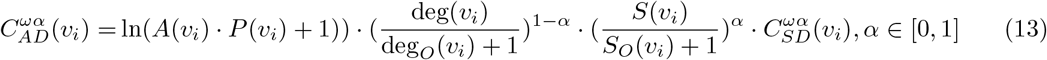

*A*(*v*_*i*_) and *P* (*v*_*i*_) (*A*(*v*_*i*_) ≥ 1, *P* (*v*_*i*_) ≥ 1) in the first term of the equation (Eqn. (13)) correspond to the number of authors and the number of publications of the institution. The second and third terms in the above equation (Eqn. (13)) are motivated by the form of the E-I index (Eqn. (6)), where we divide the total degree and the total strength (similar to E+I) by the internal cooperation degree and the internal cooperation strength (similar to I), respectively. In order to have a positive centrality score for all institutions, we added one to the first term and the denominators.

## 3 Results

### 3.1 Publication landscape

We examined trends in the number of publications in the field over the last two decades and the number of institutions involved.

Additional file Fig. 1(a) shows the number of publications on MM in SSA in 2002-2022. Considering that the information used in this study was collected in June 2022, it can be stated with certainty that the number of publications on this topic has steadily increased.

**Figure 1.**
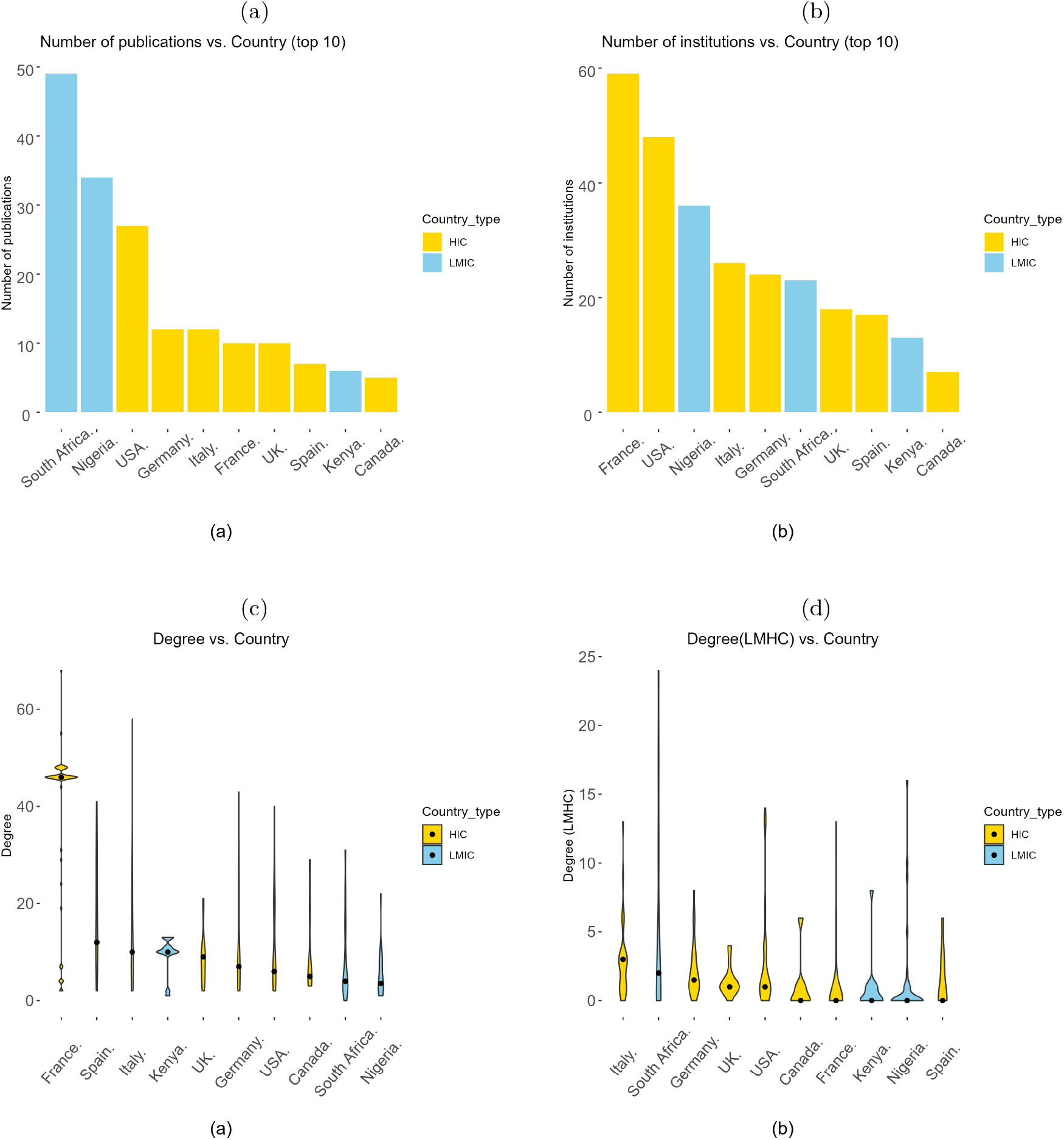
Country-level. (a) Bar charts of the number of publications and (b) Bar charts of the number of participating institutions in this field by country (top 10) between 2002 and 2022. (HICs in yellow. LMICs in blue) (c) Violin plot[39] of the degree of institutions in different countries and (d) Violin plot of LMHC degrees of institutions in different countries. (The black point is the median.) The total degree indicates the extensiveness of the collaboration and LMHC degree indicates the extensiveness of the CILC. The violin plots depict distributions of the degree of institutions from the ten countries using density curves.

Additional file Fig. 1(b) shows a histogram of the number of institutions involved in each publication. Over 50% of publications are written by authors from a single institution, and nearly 80% of publications were authored in collaboration with no more than four different institutions.

### 3.2 Country level

We compared the main metrics for different countries and then investigated the collaboration network at the country level.

In Fig. 1, We present the number of publications and contributing institutions by country. Over the past two decades, researchers from South Africa and Nigeria have been the most active, followed by HICs like the USA, Germany, Italy, the UK, France, and Spain, along with Kenya and Canada. It is important to note that South Africa, Nigeria, and Kenya are all local economic powers (second, first and fourth in SSA in terms of GDP in 2022[38], respectively).

Fig. 1(b) shows that the country with the most institutions involved is France with nearly 60 different institutions, followed by the United States with nearly 50 different institutions. The next countries are Nigeria Italy, Germany and South Africa. This is followed by the United Kingdom, Spain and Kenya with more than 10 institutions.

Fig. 1(a) and 1(b) show that while South Africa ranks sixth in the number of institutions, it leads in publications. Conversely, France, despite having the most institutions involved, ranks sixth in publications, suggesting strong internal collaboration among French institutions. Institutions in LMICs, especially South Africa and Nigeria, contribute significantly to publications, reflecting their regional influence in studying MM. However, HICs like France and the United States have more institutions participating, indicating broader collaborations.

Fig. 1(c) and (d) show the distributions of degrees by country, indicating the propensity of institutions in different countries to cooperate. Fig. 1(c) shows the total degree, while Fig. 1(d) shows the degree for the LMHC bipartite graph. The median degree of French institutions is significantly higher than others, and the majority of French institutions have a degree above 30, with only a few smaller values below 10, but the median LMHC degree of French institutions is 0, indicating that there is currently not much cooperation with institutions in LMICs. Among the HICs, some institutions in Italy and Germany tend to cooperate with institutions in LMICs. Among the LMICs, some South African institutions tend to collaborate across income levels. The complete country-level collaboration network can be found in the Additional file Fig. 2. As this study focuses on LMICs, the bipartite graph is shown in Fig. 2. The USA has extensive cooperation with LMICs, including South Africa, Nigeria, and Kenya in SSA, as well as Egypt, and Iran. Other HICs, such as Switzerland, Israel, Greece and the Netherlands, also collaborate widely with LMICs. South Africa, Nigeria, Algeria and Brazil maintain strong ties with HICs. However, there is noticeable polarization within SSA: South Africa and Nigeria are leaders in research and CILC, while countries like the Democratic Republic of the Congo, Tanzania, Ghana, and Botswana have limited collaborations with selected HICs.

**Figure 2.**
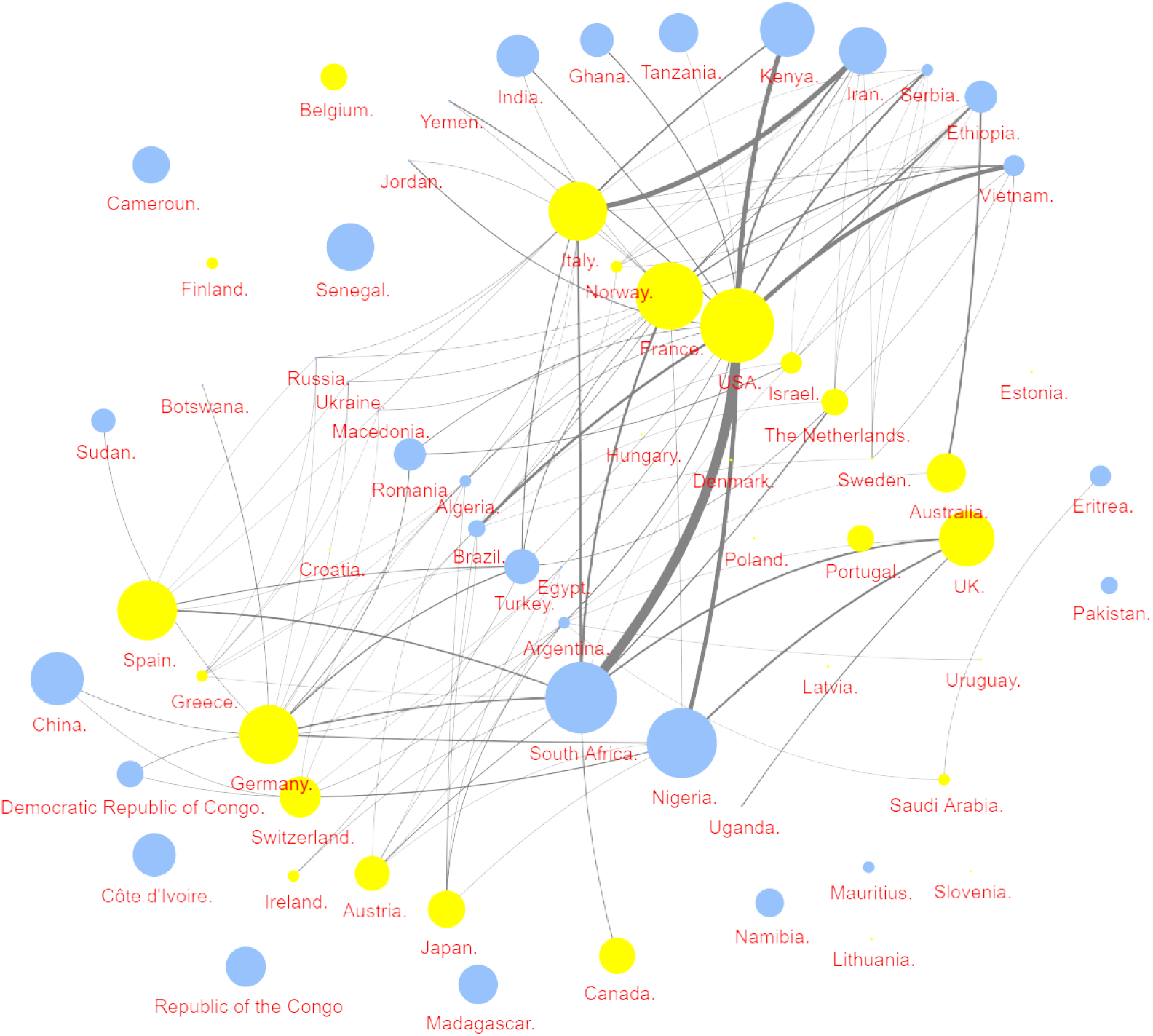
Country level CILC graph. This graph focuses on cross-income-level collaborations of institutions in LMICs with institutions in HICs. LMICs in blue, HICs in yellow. The size of the nodes is proportional to the number of authors and the thickness of the edges is proportional to the strength of the collaboration.

### 3.3 Institution level

We first analyzed the E-I index distribution of the institutions and then listed institutions leading in the metrics we proposed. We also investigated collaboration networks at the institutional level. From a holistic perspective, the original E-I index (Equation.6) is 0.2226 for the LMICs’ institutions group and -0.6034 for the HICs’ institutions group. There is slightly more external collaboration than internal collaboration in LMICs. In contrast, HICs mostly collaborate internally. The Income level regionalisation index (Equation.7) is -0.3199 for the LMIC’s institution group. This suggests that although there is more external collaboration in LMICs, the strength of internal collaboration is higher. The strength E-I index is -0.6702 for the HIC’s institutions group, which is close to the original E-I index.

For each institution, as shown in Fig. 3(a), internal collaborations dominate when the E-I index is between -0.7 and -0.5, with significantly more HIC institutions than LMICs. However, some LMIC institutions exceed HICs in external collaboration when the index is above 0.5. The KolmogorovSmirnov test shows a significant difference between the E-I index distributions of HICs and LMICs. Additional file Fig. 3 shows the leading institutions in the number of publications in this field.

**Figure 3.**
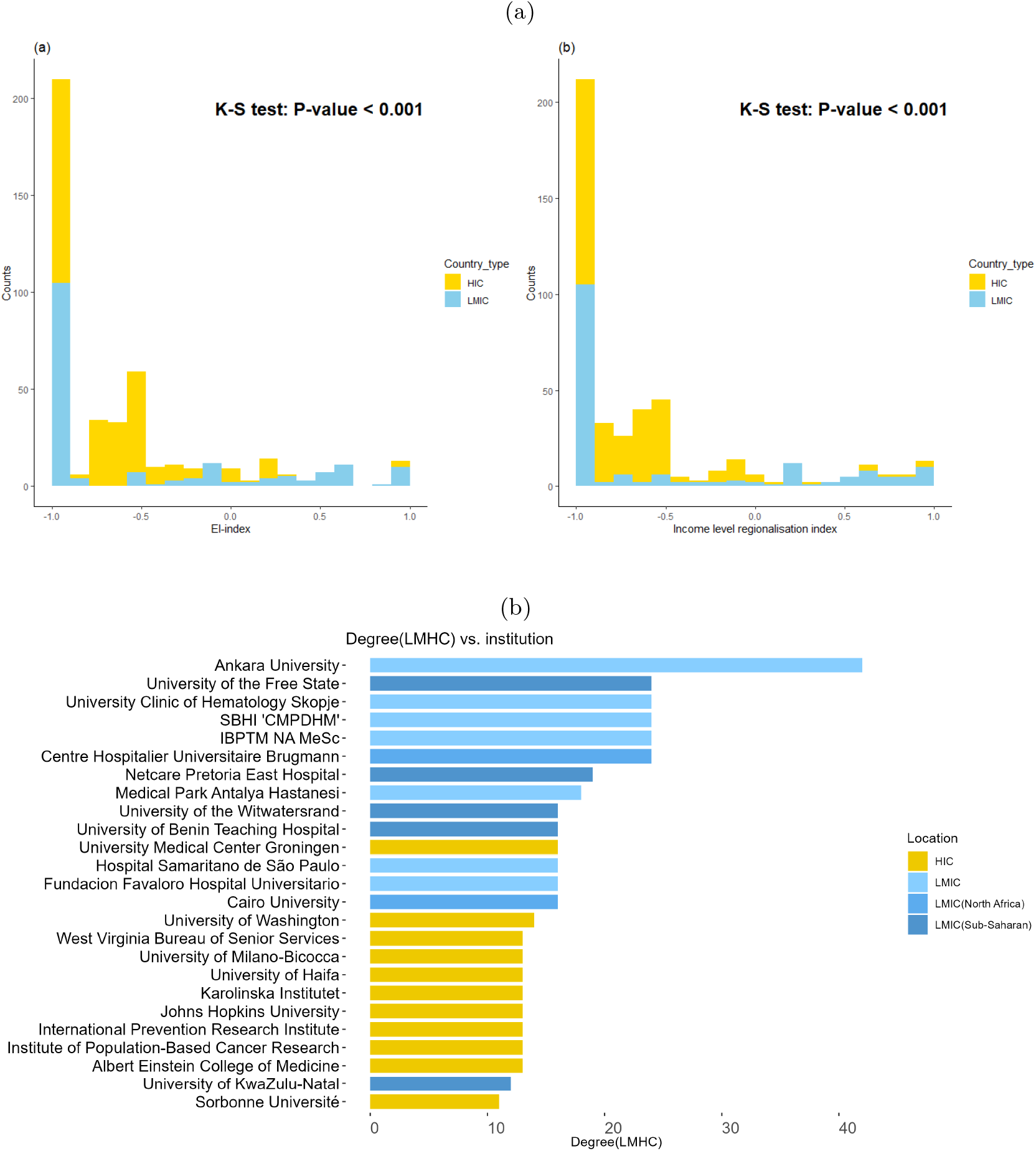
Institution-level CILC metrics. (a) Stack histograms for institutional original E-I index and institutional Income level regionalisation index. (b) Collaborations between institutions from HICs and institutions from LMICs: LMHC degree (the extensiveness of the CILC) of institutions (top 25) between 2002 and 2022.(‘SBHI “CMPDHM”‘ is short for ‘State Budgetary Healthcare Institution “Center of Medical Prevention of Department of Health of Moscow”‘, ‘IBPTM NA MeSc’ is short for ‘Institute of Blood Pathology and Transfusion Medicine of the National Academy of Medical Sciences of Ukraine’)

Sixteen of the top 26 institutions are in SSA, with South Africa (6), Nigeria (4), and Kenya (4) leading. Only ten HIC institutions are represented: three from the USA and two from Germany. The University of Cape Town leads with 12 publications, followed by the University of Witwatersrand and the University of Benin Teaching Hospital with seven each. Overall, institutions in South Africa and Nigeria, two large countries located in SSA, have contributed more publications in the field than any other country. institutions from several HICs outside the region, notably the USA and Germany have made a greater contribution in this area.

Additional file Fig. 4 shows the number of authors involved in publications from a given institution. Six of the top 24 institutions are located in the United States, five in South Africa, three in Nigeria and two in China. Specifically, over 40 researchers are affiliated with the University of Cape Town in South Africa. This is followed by the Hospital Clinic Barcelona in Spain with around 25 authors, and the University of Witwatersrand and the Fred Hutchinson Cancer Research Center in the United States. In general, institutions from South Africa are most prominent among those from SSA, while institutions from the United States and Spain are most prominent among those from HICs.

**Figure 4.**
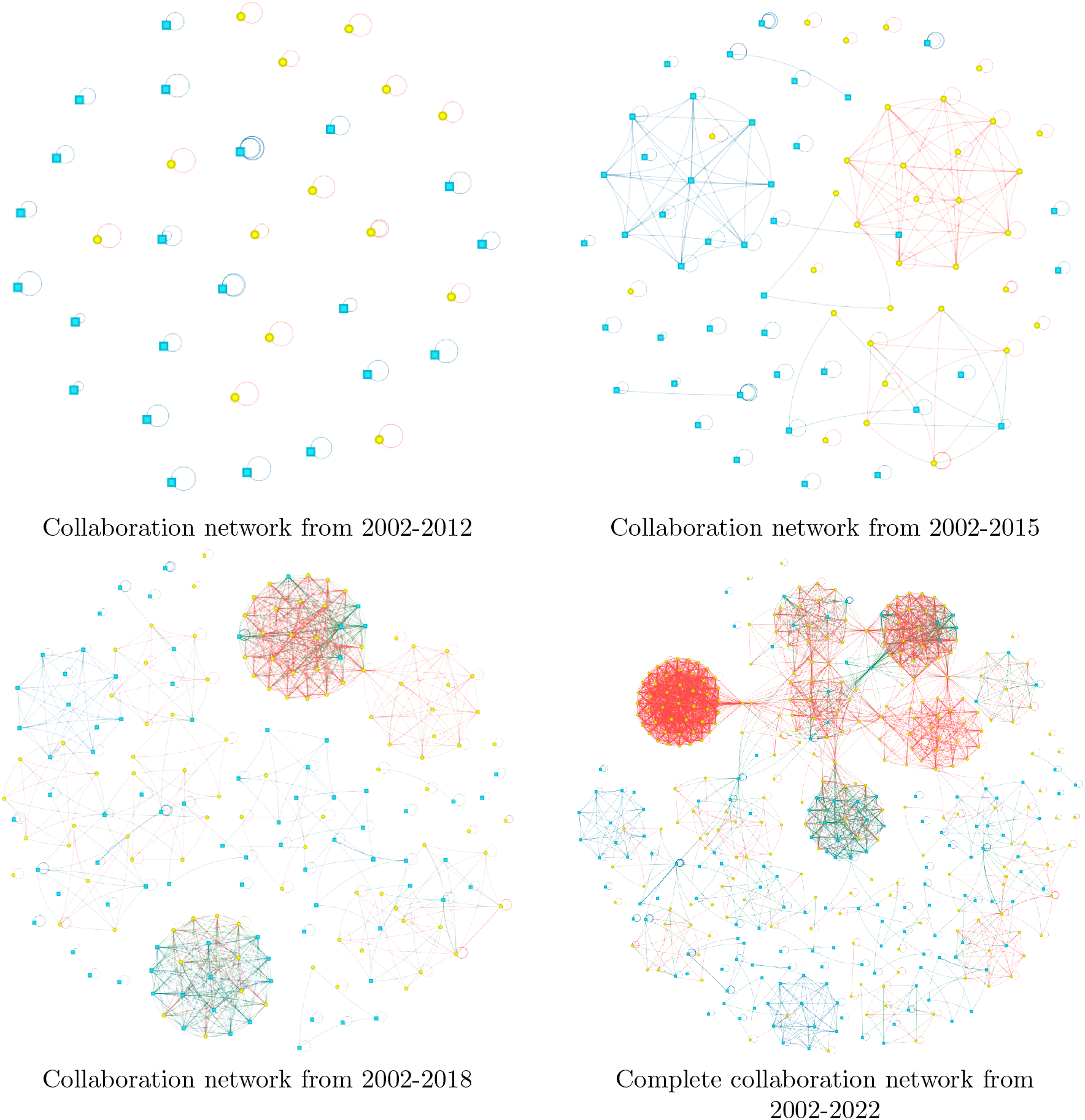
Institution-level collaboration network evolution. Vertex colours: Institutions from LMICs in blue, Institutions from HICs in yellow; Edge colours: LMLM (Intra-group collaboration among institutions in LMICs) in blue, LMHC (CILC) in green, HCHC (Intra-group collaboration among institutions in HICs) in red.

Fig. 3(b) highlights institutions likely to collaborate across income levels. LMIC institutions lead in CILC, with Ankara University in Turkey having the most collaborations with HICs. The University of the Free State in South Africa and several others, including institutions in Macedonia, Russia, Algeria, and Ukraine, follow with around 25 collaborators each. Eleven of the top 26 are in HICs, four of which are in the USA. Institutions in Turkey and South Africa, among LMICs, have the most CILC with HICs.

Fig. 4 outlines the evolution of institutional collaboration networks across four stages. From 2002 to 2012, publications were solely produced by individual institutions, predominantly in LMICs, with no collaboration. In 2013-2015, inter-institutional collaborations emerged, mainly within the same income regions. Between 2016-2018, the number of contributing institutions grew, with increased inter-institutional and CILC. The 2019-2022 period saw rapid growth in collaborations, a rise in achievements, and the formation of more closely-knit research communities. These general evolutions coincide with the trend of the publications (Fig. 1(a) in the additional file).

Fig. 5 in the Additional File shows the complete collaboration network at the institution level in high resolution. Fig. 6 in the Additional File is the subnetwork constructed in the same way but only for publications explicitly tagged as ‘clinical trial’ and ‘observational study’ on PubMed.

**Figure 5.**
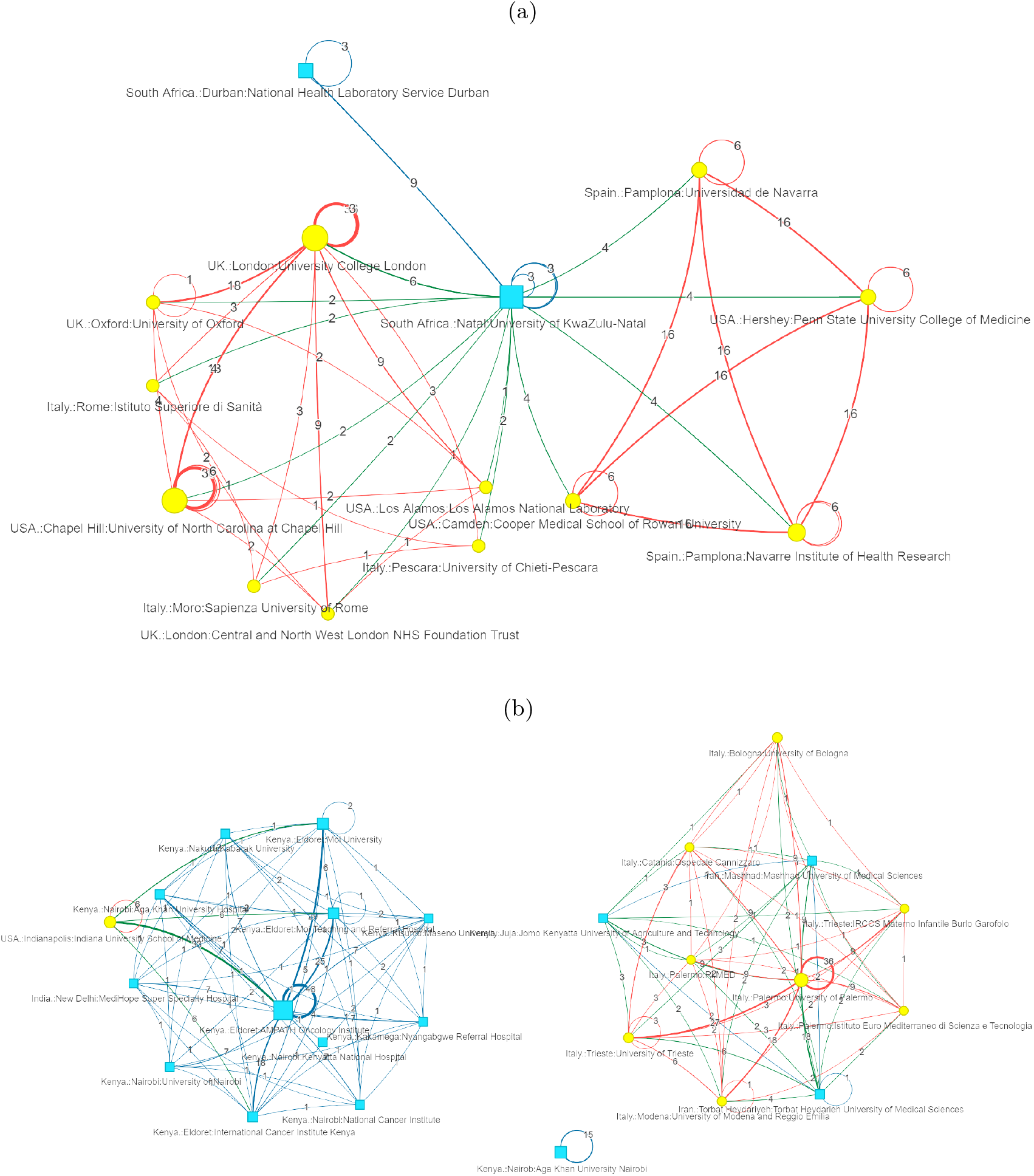
Keyword-search subnetworks. (a) Network graph centred on the University of Kwazulu-Natal. (b) Network graph of institutions in Kenya. Vertex colours: Institutions from LMICs in blue, Institutions from HICs in yellow; Edge colours: LMLM (Intra-group collaboration among institutions in LMICs) in blue, LMHC (CILC) in green, HCHC (Intra-group collaboration among institutions in HICs) in red. The number of contributions from affiliated authors determines the node size and the strength of collaboration determines the thickness of the edge

#### 3.3.1 The University of Kwazulu-Natal in South Africa

In the interactive tool, the network can be analysed further. In addition, the search function allows one to view sub-graphs of the network based on user-specified institutions or countries.

The University of KwaZulu-Natal in South Africa is a world-renowned institution. Its uniqueness of being primarily active in CILC is fully revealed with the help of interactive tools. The sub-network graph was derived from a search for the exact keyword ‘KwaZulu-Natal’. Thus, the network is centred on that university with neighbours.

In Fig. 5(a), we performed a representative analysis of a sub-network centred by ‘the University of Kwazulu-Natal’. In terms of collaboration with institutions from LMICs, we found only one collaboration with the National Health Laboratory Service in Durban, a South African national government institution, which has the highest intensity of collaboration in the network. However, the university has collaborated with several institutions in HICs including University College Lon-don, Cooper Medical School of Rowan University in the USA and the University of Navarra in Spain.

#### 3.3.2 Institutions in Kenya

Our tool also allows one to search for countries or cities. The network will then show individual sub-graphs with the institutions in that country or city as the core and combine them into a single graph.

Kenya, as one of the top economies in SSA after Nigeria and South Africa[38], domestic institutions are far inferior to those in South Africa and Nigeria in terms of the breadth of CILC, thus presenting a representative and interesting pattern of cooperation. The sub-network graph was derived from a search for the exact keyword ‘Kenya’. It includes all Kenyan institutions, as well as the neighbours.

Fig. 5(b) shows the network is divided into three clusters: the cluster on the top consists of individual institutions, the cluster on the right consists mainly of Kenyan institutions, and the cluster on the left consists mainly of Italian institutions.

In the Kenya-dominated cluster, the largest node is the AMPATH Oncology Institute, which, along with the Moi Teaching and Referral Hospital and Moi University, shows strong internal collaboration. Notably, this cluster includes two foreign institutions: Max Super Specialty Hospital in India and Indiana University School of Medicine in the USA. This US institution collaborates with four Eldoret institutions in Kenya, including the three mentioned above, as well as the International Cancer Institute, and the strength of collaboration is among the higher values in this cluster.

In the Italy-dominated cluster, the Jomo Kenyatta University of Agriculture and Technology from Kenya has low cooperation with other cluster members. This cluster also includes two universities from Iran. The largest node is the University of Palermo, which has a high intensity of collaboration with several Italian institutions.

#### 3.3.3 Betweenness centrality, eigenvector centrality and adjusted degree centrality ranking of institutions

We used Python NetworkX to implement the betweenness centrality and eigenvector centrality algorithms and the top 20 institutions ranked by the algorithms were highlighted in the network graph using Pyvis. The leading institutions identified by betweenness centrality (Figure.7 in Additional file) are indeed the hub institutions between the communities, Whereas the leading institutions identified by eigenvector centrality (Figure.8 in Additional file) are the majority of institutions in an intermediate community within several large communities with good connectivity.

Using the adjusted degree centrality function (Eqn. (13)), the results of the sorting for different values of *α* are shown in the Additional file Table.1.

Table 2 shows the main metrics of the top 15 institutions when *α* = 0.5. This value for the tuning parameter implies that equal weight is given to the breadth and strength of the institution’s CILC. Compared to the original algorithm, the adjusted algorithm gives better results. (detailed discussion can be found in the Additional file Section.7)

**Table 2:**
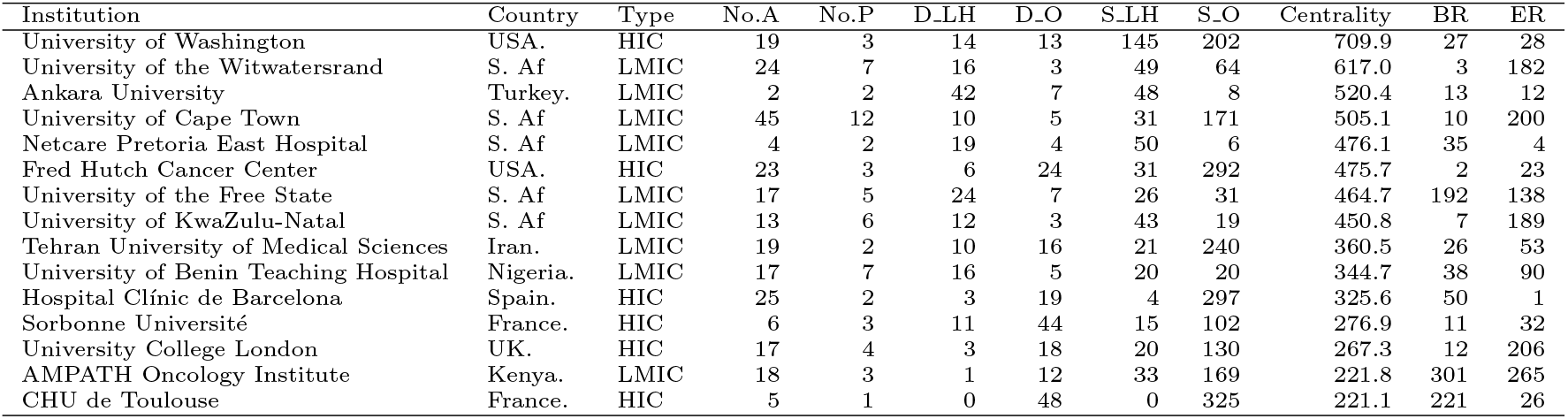
Main metrics of the top 15 institutions at *α*=0.5, (S. Af is short for South Africa, BR is short for betweenness centrality ranking, ER is short for eigenvector centrality ranking)

It is necessary to rank the local institutions located in SSA separately, When *α* is 0.5, the top 15 rankings can be found in the additional file Table 2. Seven of the top 15 institutions are located in South Africa, while four are in Nigeria, three are in Kenya, and one is in Ethiopia. These countries are in SSA. Most top institutions are universities, but the highest-ranked is the University of the Witwatersrand in South Africa. The Netcare Pretoria East Hospital ranked third could be a potential partner for clinical trials. The AMPATH Oncology Institute in Kenya (ranked seventh) and Echolab Radiology and Lab Services in Nigeria (ranked fourteenth) stand out as unique research-focused institutions.

## 4 Discussion

In this study, we constructed weighted networks based on publications to identify the countries and institutions active in CILC in the field of MM in SSA, which could provide relevant scientific information for strategic planning of health organizations[40].

The increase in publications on MM in SSA and the network evolution have shown clear growth in this field. The E-I index, expanded indicators, and network patterns show that internal cooperation in this area is currently dominant, especially among HICs. There is great potential for beneficial CILC, which may allow this interest to be structured more effectively.

Combining indicators such as the number of papers published, the number of authors, and the breadth of collaboration, the HICs leading in this area of research are the United States, France, Italy and Germany. Among LMICs, the leaders are South Africa, Nigeria and Kenya, all of which are regional powers located in SSA. In comparison, HICs lead in terms of the number of institutions and the breadth of collaboration, while LMICs have prominence in terms of the number of papers published. We found that many institutions in LMICs in SSA have only self-cooperation, such as Cote d’Ivoire, Senegal, Cameroon, the Republic of Congo and Namibia, and these institutions may benefit from CILC in the future.

The University of Kwazulu-Natal in South Africa works closely with institutions in HICs such as the USA, Spain and the UK, but not with institutions in other LMICs. This interesting pattern might make it a potential future pivotal institution in SSA. More focused and detailed work on this representative pattern could be undertaken.

Our analysis of Kenyan institutions revealed two main clusters. One cluster is dominated by Kenyan institutions, the centre of which is the AMPATH Oncology Institute in Eldoret, and the other is dominated by Italian institutions, of which Jomo Kenyatta University of Agriculture and Technology is the only Kenyan institution.

In the centrality algorithm of institutions section, the betweenness and eigenvector centrality tend to identify key institutions within or between large communities where internal collaboration between institutions in HICs is dominant, and these results did not capture CILC between institutions in HICs and those in LMICs. Therefore, an adjusted degree centrality algorithm was used to rank the CILC performance of the institutions. The main indicators of the algorithm were the number of publications, the number of authors, the total number of degrees, the degree of the LMHC category, total collaboration strength and the collaboration strength of the LMHC category. Those institutions engaged in extensive and strong CILC are ranked higher under the algorithm. The results are also consistent with the comparative analysis of the main metrics. The local universities in the table could measure the universities in Africa in terms of research output and global rankings[41]. It is interesting to note that most leading institutions are universities in various countries or regions. Considering the specificity of the medical research field, hospitals among the leading institutions may be unique collaborators for cooperation in conducting clinical research on African patients with MM.

Our tool enables country, city, and institution-specific keyword searches with optional group colouring and size functions proportional to author count and collaboration strength. This allows for a quick, intuitive comparison of collaboration gaps across HICs and LMICs. In addition, the collaboration network graph drawn after searching for specific institution name filters can also provide evidence for the adjusted degree centrality algorithm in our paper. A time slider helps analyze the network’s evolution. While data cleaning and network construction were challenging, our scalable code and expanded keyword-matching dictionary will streamline future work. Extending these tools to large-scale clinical research networks is a promising idea.

### 4.1 Potential implication for policy and practice

As highlighted by Sambo and colleagues, the key cancer prevention and control challenges in SSA include insufficient recent and comprehensive data for cancer and death registration, inadequate or no information about cancer, insufficient numbers of skilled healthcare personnel, and scarce local, effective, and sustainable research and absence of collaboration or coordination of interventions in stakeholders and donors to combat cancer.[42]

Although there has been a significant increase in collaboration and engagement in MM in SSA over the past few years, our findings suggest that collaborations between local institutions are insufficient. Initiatives that increase incentives and opportunities for regional collaborations, as well as enable access to necessary resources such as journal content and funding, would serve to strengthen the regional network and build capacity for regionally relevant research. Ideally, capacity-building research programmes on MM in SSA can be started from the leading local countries and institutions or the ones that have already collaborated with national government institutions, such as the University of Kwazulu-Nata. Besides, the involvement of the local hospitals can promote data collection and clinical trial research. A great paradigm is the Men of African Descent and Carcinoma of the Prostate (MADCaP) consortium, which presently includes multiple centres representing populations in Senegal, Ghana, Nigeria, Sudan, Uganda, Botswana, and South Africa, aims to improve the prevention, screening, and treatment of cancer in affected communities, and the quality of life of men who have been diagnosed with cancer. [43]

Secondly, CILC is still lacking in this area, Institutions actively seeking CILC may bring external funding support to the local area, and the construction of a research network may inform the allocation of resources in favour of equitable allocation of funds [44]. A stronger CILC can further the training of professionals in research skills as well as the exchange of experiences to determine shared research priorities. Interaction between universities and the local hospitals can be essential in securing joint ownership of research, and critical for sustainable research development in which health practitioners can innovate care based on research findings to improve population health. [45] Taking into account that CILC is often influenced by external pressures, such as policy constraints. The leading institutions identified in this paper imply their openness to, and feasibility of, CILC, and thus they may be potential partners. Our network analysis methods and the interactive tool are transferable to other domains, providing researchers with information on active institutional collaborations that contribute to global health.

### 4.2 Limitations

A large proportion of contemporary academic publications from English-speaking HICs[19], a circumstance that may create a natural research affinity toward Anglophone regions in SSA[46]. In addition, the persistent under-representation of local authors in academic publications [47] is a challenge to identify local experts, and our study partially addresses this issue by identifying those local institutions whose authors strongly collaborate.

The dataset used in this article was obtained from Pubmed databases, it cannot be guaranteed that it includes all relevant publications, and the publications are not necessarily focused on MM, they could be about several cancers including MM. As a result, the analysis that was conducted using this dataset might not be all-encompassing. However, according to Falagas and colleagues[48], who compared PubMed, Scopus, Web Of Science and Google Scholar in their paper, PubMed is the optimal choice in terms of scientific database[49].

Moreover, PubMed’s affiliation data is often inconsistent, with occasional missing information. Some publications only list the first author’s institution, omitting others. Manual checks revealed that omitted co-authors are from the same institution as the first author, so they were assigned accordingly during data processing, though this method may introduce bias.

During the curation of the dataset, systematic difficulties were encountered: The institutions and affiliations change dynamically. For example, the University of KwaZulu-Natal (UKZN) was established on 1 January 2004 following the merger of the University of Natal and the University of Durban-Westville.[50] A 2003 publication in which the authors were affiliated with the University of Natal was therefore manually assigned to UKZN, We used ROR API to standardize but some institutions, especially in less represented regions, e.g. the hospitals or national institutions in some sub-Saharan countries such as Kenya (International Cancer Institute Eldoret, Moi Teaching and Referral Hospital) and Eritrea (Orotta National Referral Hospital, Ghindae Zonal Referral Hospital and National Health Laboratory Eritrea) etc., could not be identified correctly by ROR, but ROR would always match them to registered institutions, thus may introducing bias although we manually checked.

The strength of collaboration is measured by the product of the number of authors from each institution, assuming collaboration between every pair of authors. For authors with multiple affiliations, it’s assumed these institutions collaborate, which may also introduce bias. For sub-networks of multiple institutions that have been working together for a long time, there may be a higher chance that some of the authors are affiliated with several of the involved institutions. Our results may therefore inflate the strength of these collaborations. In our data set, a collaboration between AMPATH Oncology Institute, the Indiana University School of Medicine, Moi University, and Moi Teaching and Referral Hospital is an example of such a strong and longstanding collaboration.

The interactive tool is only accessible by cloning from the GitLab repository and running it locally. It requires special data processing before importing. The tool automatically generates network graphs based on filtering, but users can only drag nodes to adjust positions, making it less user-friendly. Additionally, the displayed information is limited; hovering over an institution’s node shows the PMID and author list but without detailed correspondences.

## 5 Conclusions

In general, for institutions studying MM in SSA, there is relatively little CILC and a clear centrality, i.e. institutions in HICs tend to collaborate with only a few local institutions in South Africa and Nigeria. The leading local institutions in SSA do not collaborate much with other local institutions. We notice this trend and use metrics to highlight it to give some validation across patterns. We adjusted the degree centrality algorithm in the weighted network, i.e., the degree centrality score was adjusted using the E-I index to identify institutions in the field that are active in CILC. The central institutions identified contribute to the stability of the network, help connect institutions and serve as conduits of knowledge, resources and expertise[51].

In addition, the effect of using network analysis for this study is clear, and the network visualization interaction tool may be a useful tool in the future. With the help of the interactive tool, the stakeholders can visualise the information about collaboration between institutions in the field. This paper provides some reference and assistance in promoting such cooperation.

## Data Availability

All data produced in the present study are available upon reasonable request to the authors

https://gitlab.bham.ac.uk/spillf-systems-mechanobiology-health-disease/collaboration-network-multiple-myeloma

## Acknowledgements

We are grateful to Natanya Jennings and Sue Bailey from Bristol Myers Squibb for sharing ideas and providing valuable feedback on the results.

## Funding

F.S. was supported by a UKRI Future Leaders Fellowship, grant no. (MR/T043571/1).

### Abbreviations

MM: multiple myeloma
SSA: Sub-saharan Africa
LMIC: Low-and-middle-income country
HIC: High-income country
CILC: cross-income-level collaboration

## Availability of data and materials

Please contact the corresponding author for data and material requests. The source code for the interactive tool can be found on the main branch on our Gitlab page https://gitlab.bham.ac.uk/spillf-systems-mechanobiology-health-disease/collaboration-network-multiple-myeloma. The branch named Data contains the code for data collection and cleaning.

## Ethics approval and consent to participate

Not applicable

## Competing interests

The authors declare that they have no competing interests.

## Consent for publication

Not applicable

## Authors’ contributions

KY developed the code, performed the analysis and wrote the paper. SBB advised on the data crawling and cleaning and revised the manuscript. JBC and FS conceived the analytical approach and contributed to the direction and focus of the work as supervisors. FS also contributed to the methodology.

## Additional Files

### Additional file

Section.1 Publication landscape visualization

Section.2 Country-level collaboration network

Section.3 Institution-level visualization

Section.4 Subnetwork –Clinical trial and observational study publications

Section.5 Betweenness centrality and eigenvector centrality top institutions identified

Section.6 Adjusted degree centrality ranking of institutions

Section.7 Original degree centrality algorithm results and discussion

## Notes

### Competing Interest Statement

The authors have declared no competing interest.

