## Additional File for "Network analysis of cross-income-level collaboration on non-communicable disease: the example of multiple myeloma in Sub-Saharan Africa"

October 22, 2024

#### 1 Publication landscape

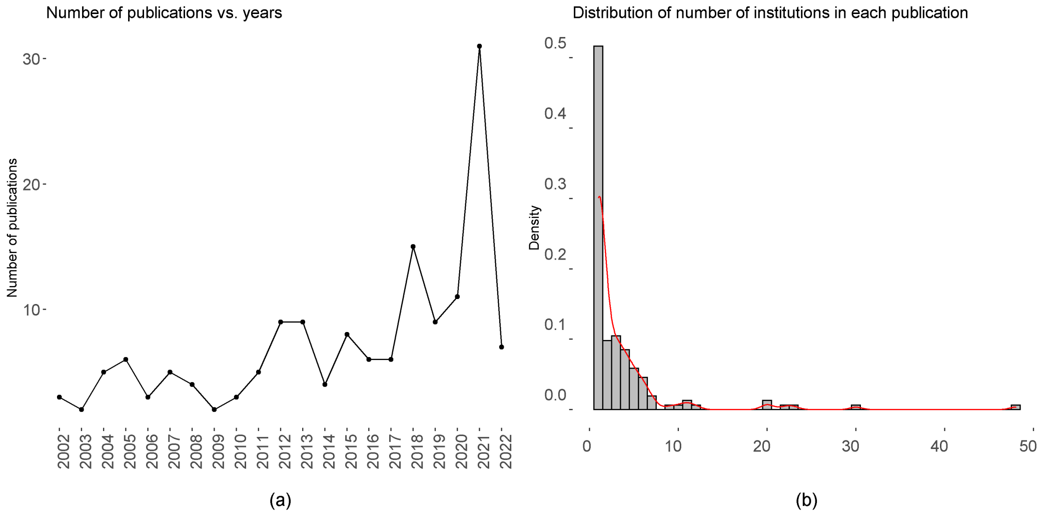


Figure 1:
**Publication statistics**(a) Line graph of the number of publications by year (as of June 2022) and (b) Histogram of the number of institutions in each publication (2002-2022).

#### 2 Country-level collaboration network

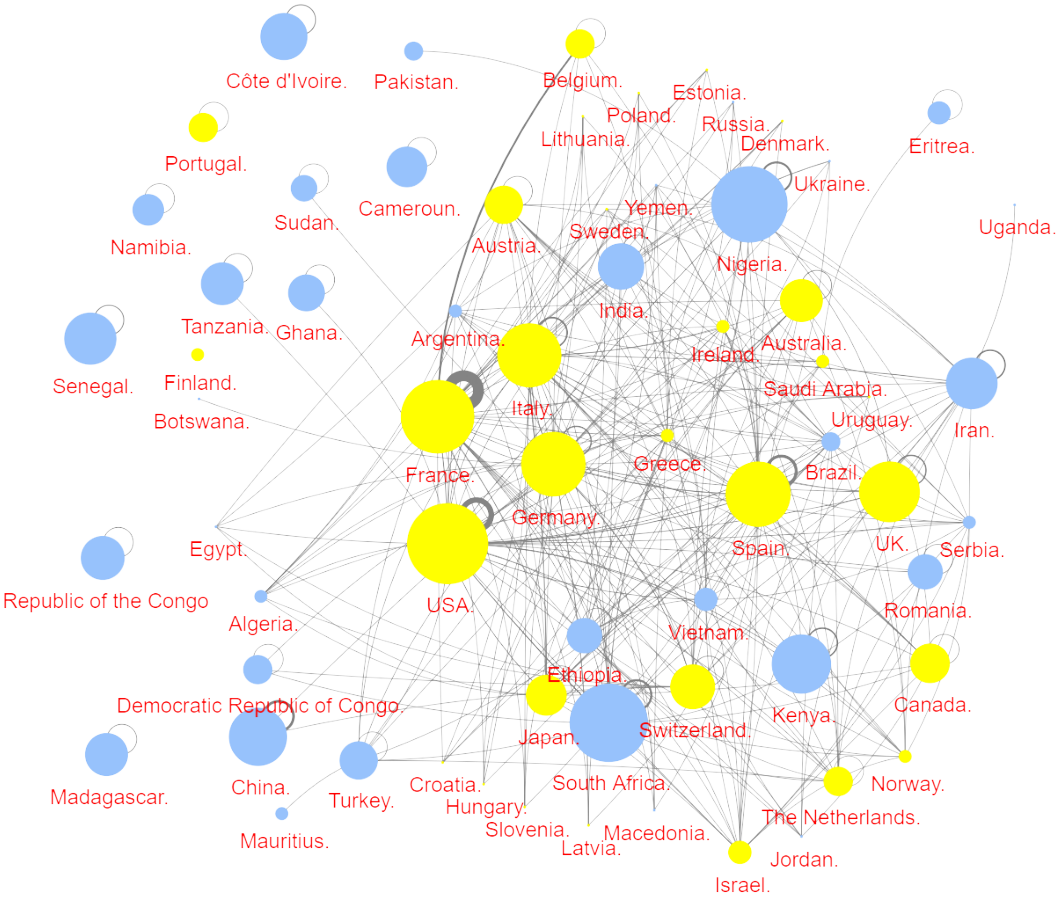


Figure 2:
**Country level collaboration network:** LMICs in blue, HICs in yellow. The size of the nodes is proportional to the number of authors in that country who have publications in the field, and the thickness of the edges is proportional to the strength of the collaboration.

### 3 Institution level

#### 3.1 Institution-level metrics

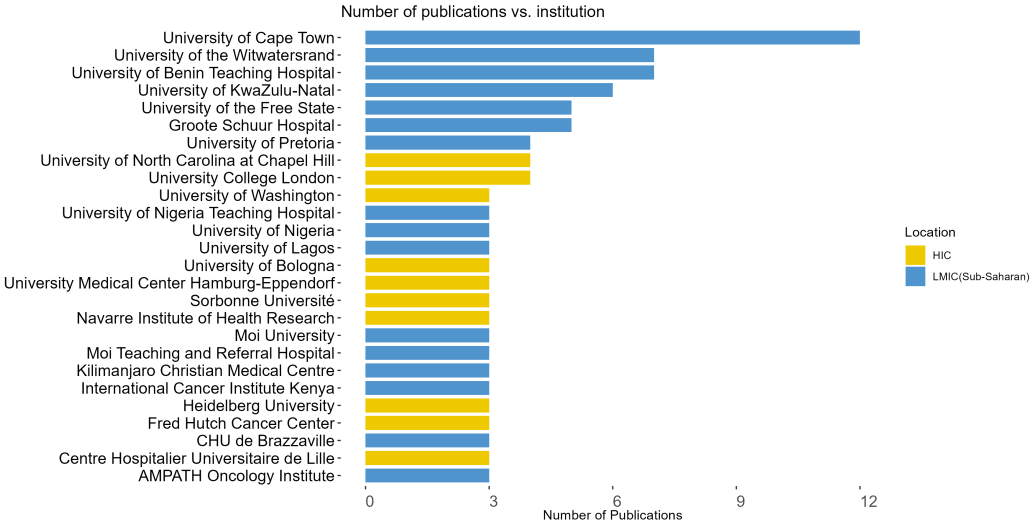


Figure 3:
**Bar charts of the number of publications on MM in SSA by institutions (top 26) between 2002 and 2022.**

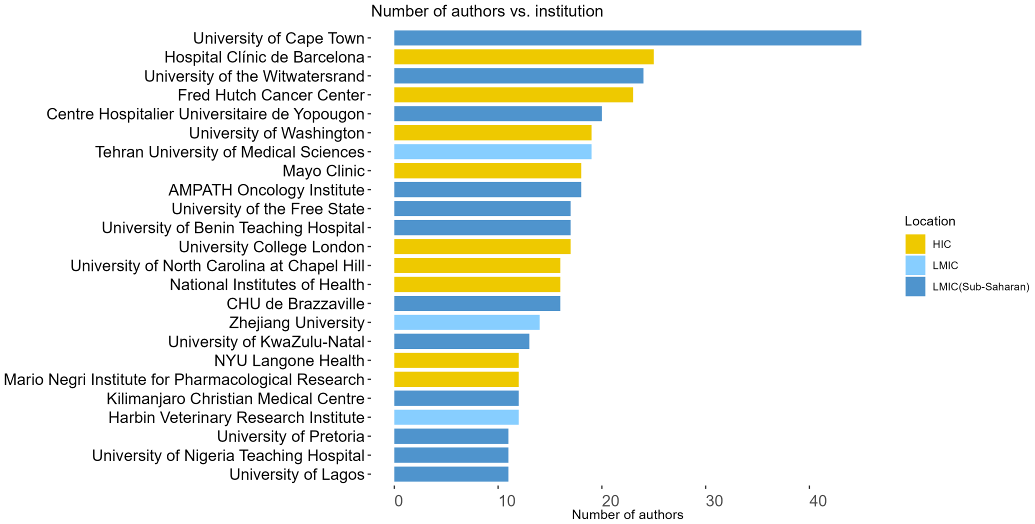


Figure 4:
**Bar charts of the number of authors by institutions (top 24).**

##### 3.2 Complete institution-level collaboration network

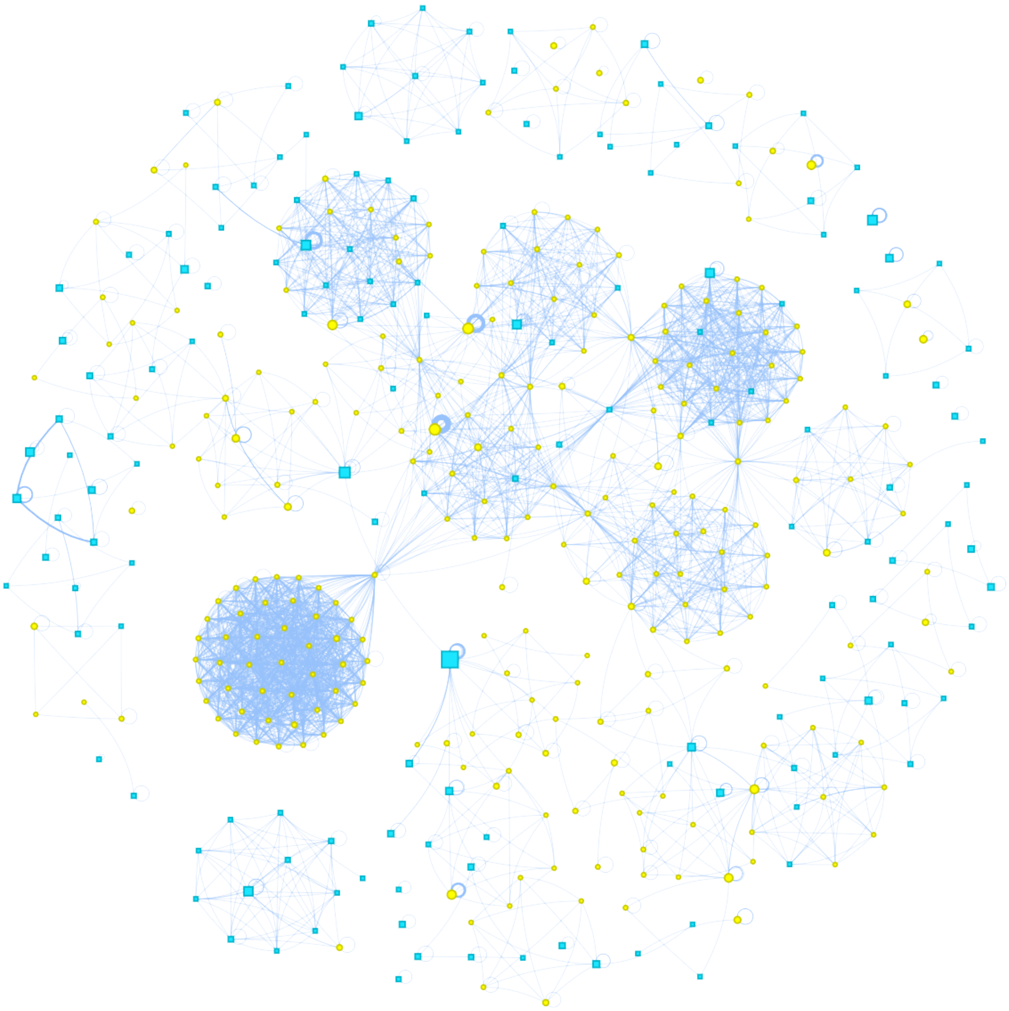

Figure 5:
**The full collaboration network at the institution level.** Institutions in HICs are in yellow, and institutions in LMICs are in blue. Node size is proportional to the LMHC degree (the extensiveness of the institution's collaboration). The thickness of the edges is proportional to the strength of collaboration.

### 4 Subnetwork –Clinical trial and observational study publications

We selected publications explicitly tagged as 'clinical trial' and 'observational study' on PubMed, seven in all. The sub-network was constructed similarly, As we can see from Fig 6, institutions from HICs were overwhelmingly dominant in this sub-network, a trend that is even more pronounced than the one shown in the full network graph. In light of this phenomenon, we scrutinised relevant publications, such as the clinical trial publication with PMId 26089396, which lists authors with affiliations from various hospitals in Spain, and finally, a large collaborative group (the appendix of this publication lists some collaborators from local hospitals in SSA), which suggests that local institutions were indeed involved in the study but their participation is limited. Among the few institutions from LMICs, only a few institutions from South Africa are local, which highlights the special role of South Africa.

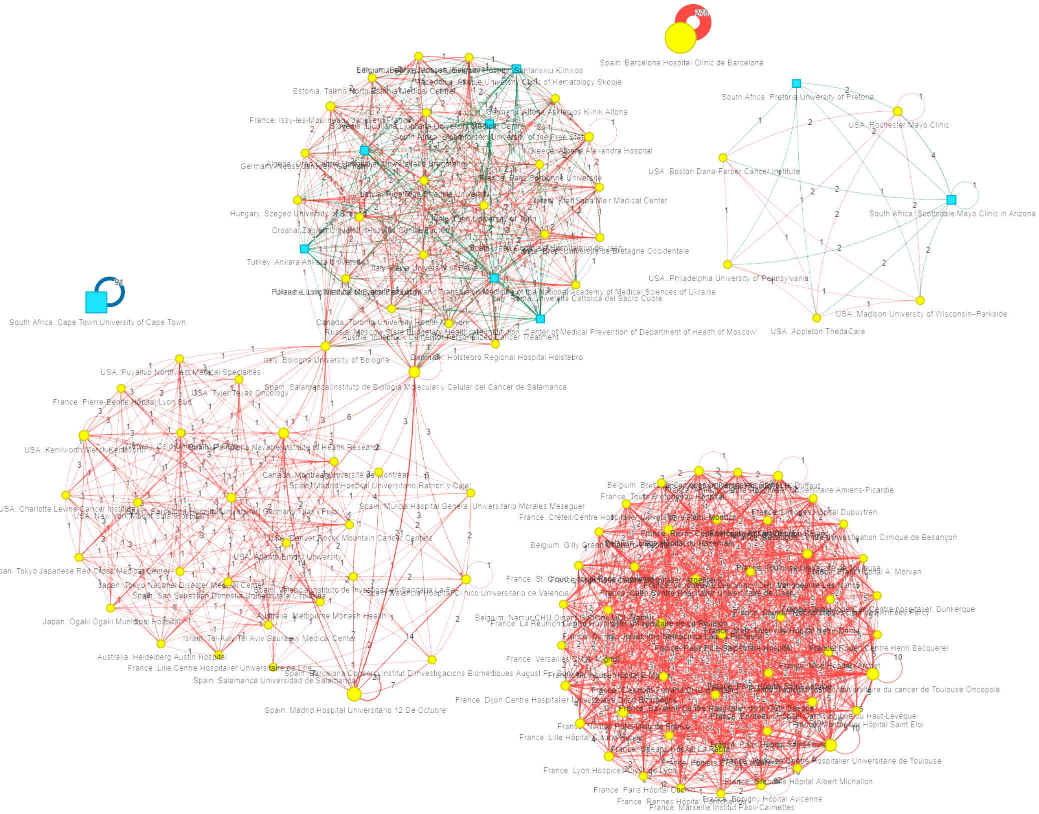


Figure 6:
**Clinical trial and observational study publications – institutional collaboration network graph** Vertex colours: Institutions from LMICs in blue, Institutions from HICs in yellow; Edge colours: LMLM (Intra-group collaboration among institutions in LMICs) in blue, LMHC (CRC) in green, HCHC (Intra-group collaboration among institutions in HICs ) in red. The number of contributions from affiliated authors determines the node size and the strength of collaboration determines the thickness of the edge

### 5 Betweenness centrality and eigenvector centrality top institutions identified

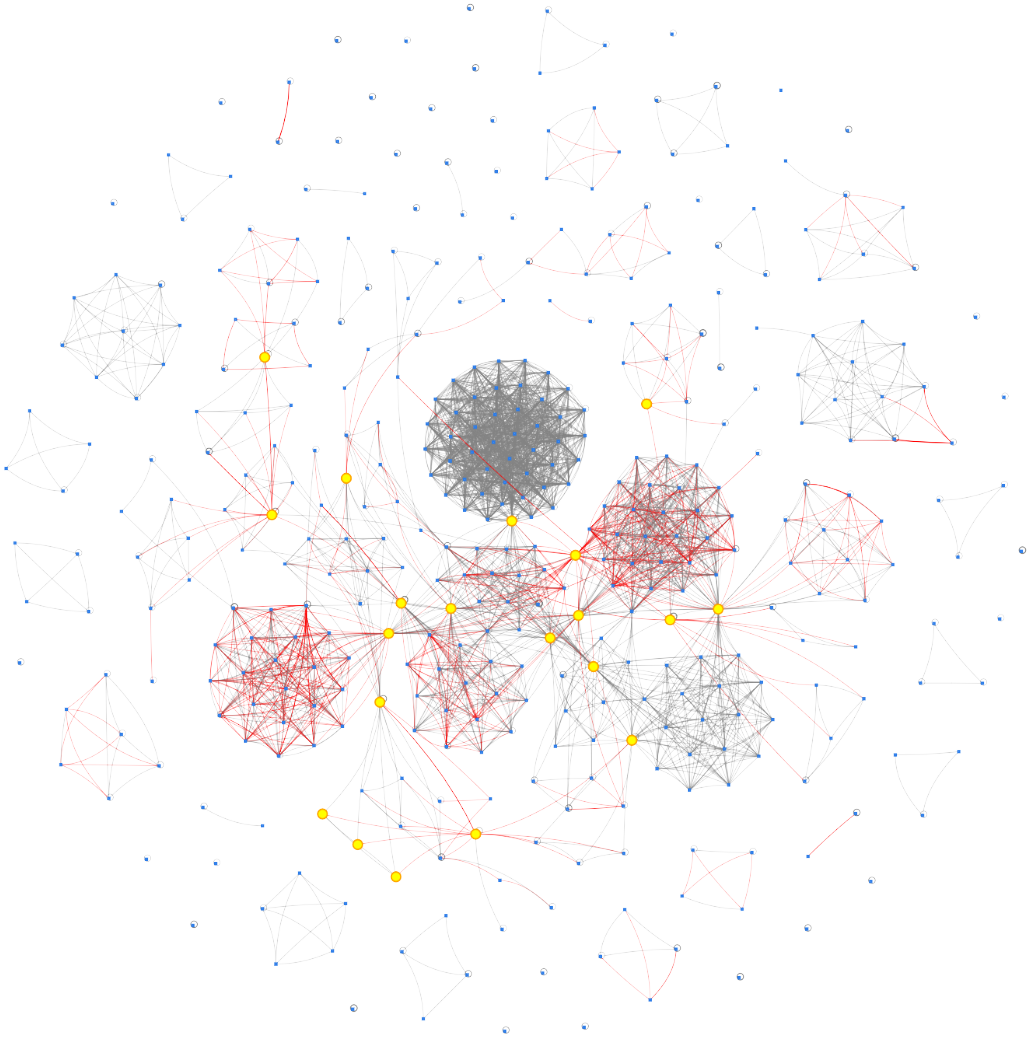

Figure 7:
**Institutional network graph.**The top 20 institutions identified by betweenness centrality are highlighted. The edges representing the cross-group collaboration are in red.

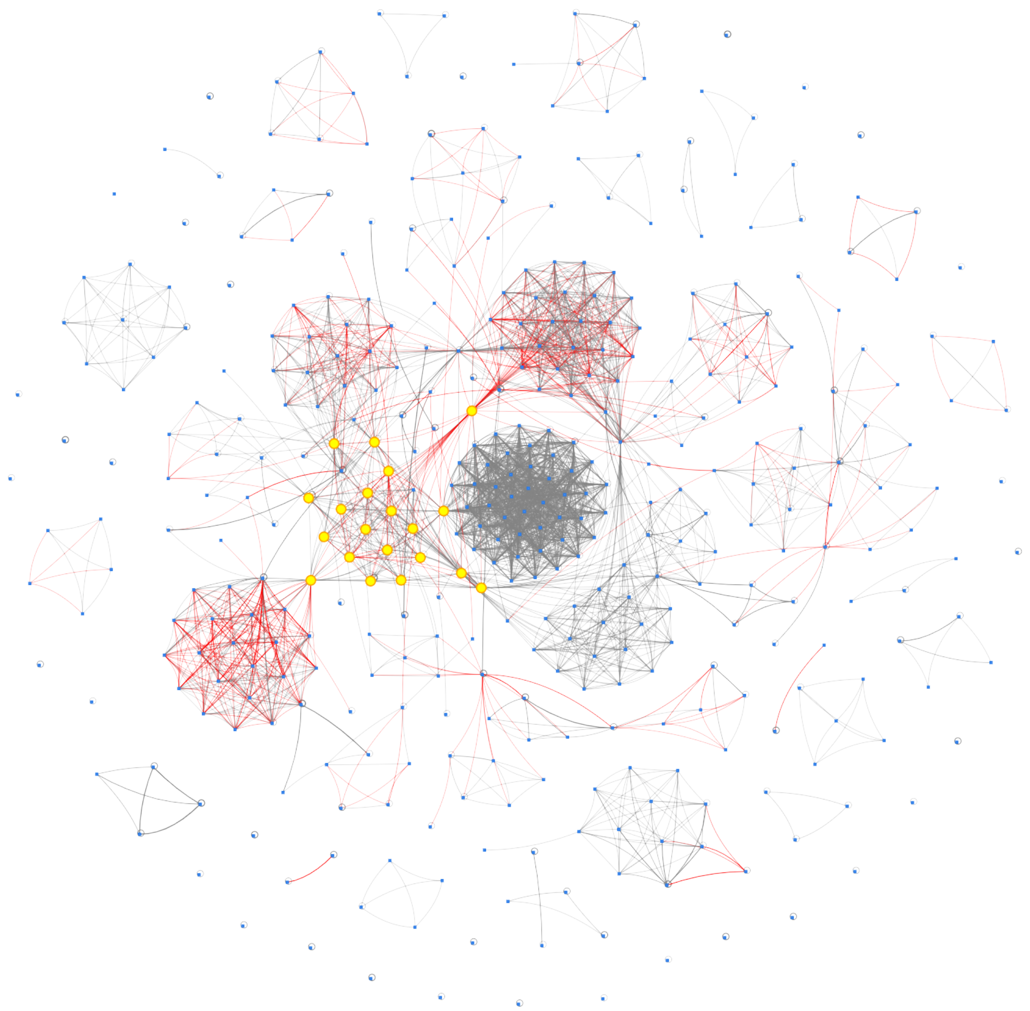

Figure 8:
**Institutional network graph.** The top 20 institutions identified by eigenvector centrality are highlighted. The edges representing the cross-group collaboration are in red.

### 6 Adjusted degree centrality ranking of institutions

The complete results of the ranking for different values of $\alpha$ are shown in Table1.

Table 1: Ranking of institutions when different tuning parameter values are used. (A smaller $\alpha$ indicates centrality is mainly determined by extensiveness, while a larger $\alpha$ indicates centrality is mainly determined by strength.)

| R | $\alpha$=0.2 | $\alpha$=0.5 | $\alpha$=0.8 |
|---|---|---|---|
| 1 | University of the Witwatersrand | University of Washington | University of Washington |
| 2 | University of the Free State | University of the Witwatersrand | Fred Hutch Cancer Center |
| 3 | Ankara University | Ankara University | University of Cape Town |
| 4 | University of Benin Teaching Hospital | University of Cape Town | University of the Witwatersrand |
| 5 | University of Washington | Netcare Pretoria East Hospital | Netcare Pretoria East Hospital |
| 6 | University of KwaZulu-Natal | Fred Hutch Cancer Center | Hospital Clínic de Barcelona |
| 7 | Netcare Pretoria East Hospital | University of the Free State | Tehran University of Medical Sciences |
| 8 | University of Cape Town | University of KwaZulu-Natal | Indiana University School of Medicine |
| 9 | Fred Hutch Cancer Center | Tehran University of Medical Sciences | University of KwaZulu-Natal |
| 10 | Sorbonne Université | University of Benin Teaching Hospital | Ankara University |
| 11 | Tehran University of Medical Sciences | Hospital Clínic de Barcelona | AMPATH Oncology Institute |
| 12 | University of Bologna | Sorbonne Université | University College London |
| 13 | Hospital Clínic de Barcelona | University College London | University of the Free State |
| 14 | Institut Paoli-Calmettes | AMPATH Oncology Institute | Centre Hospitalier Universitaire de Toulouse |
| 15 | University College London | Centre Hospitalier Universitaire de Toulouse | Institut universitaire du cancer de Toulouse Oncopole |

Rankings of the top 15 local institutions located in SSA are in Table.2.

Table 2: Main metrics of the top 15 SSA institutions at $\alpha = 0.5$

| Institution | Country | No.A | No.P | D_LH | D_O | S_LH | S_O | Centrality |
|---|---|---|---|---|---|---|---|---|
| University of the Witwatersrand | South Africa. | 24 | 7 | 16 | 3 | 49 | 64 | 617.0 |
| University of Cape Town | South Africa. | 45 | 12 | 10 | 5 | 31 | 171 | 505.1 |
| Netcare Pretoria East Hospital | South Africa. | 4 | 2 | 19 | 4 | 50 | 6 | 476.1 |
| University of the Free State | South Africa. | 17 | 5 | 24 | 7 | 26 | 31 | 464.7 |
| University of KwaZulu-Natal | South Africa. | 13 | 6 | 12 | 3 | 43 | 19 | 450.8 |
| University of Benin Teaching Hospital | Nigeria. | 17 | 7 | 16 | 5 | 20 | 20 | 344.7 |
| AMPATH Oncology Institute | Kenya. | 18 | 3 | 1 | 12 | 33 | 169 | 221.8 |
| North-West University | South Africa. | 3 | 2 | 5 | 2 | 17 | 1 | 94.8 |
| University of Nigeria | Nigeria. | 10 | 3 | 5 | 3 | 12 | 20 | 92.8 |
| Haramaya University | Ethiopia. | 2 | 1 | 10 | 14 | 42 | 29 | 86.8 |
| Moi University | Kenya. | 5 | 3 | 1 | 12 | 12 | 49 | 84.8 |
| University of Pretoria | South Africa. | 11 | 4 | 5 | 4 | 6 | 18 | 80.1 |
| Moi Teaching and Referral Hospital | Kenya. | 4 | 3 | 1 | 12 | 6 | 50 | 72.4 |
| Echolab Radiology And Laboratory Services, Benin City | Nigeria. | 1 | 1 | 9 | 0 | 11 | 0 | 68.6 |
| University of Lagos | Nigeria. | 11 | 3 | 2 | 5 | 6 | 33 | 66.5 |

### 7 Original degree centrality results

Using the degree centrality function:

$$C_D^{\omega\alpha}(v_i) = \deg(v_i)^{1-\alpha} \cdot S(v_i)^{\alpha}, \alpha \in [0, 1]$$

The results of the sorting for different values of $\alpha$ are shown in Table3.

Table 3: Ranking of institutions for different values of tuning parameter $\alpha$

| R | $\alpha$=0.2 | $\alpha$=0.5 | $\alpha$=0.8 |
|---|---|---|---|
| 1 | Institut Paoli-Calmettes. | CHU de Toulouse. | CHU de Toulouse. |
| 2 | CHU de Toulouse. | IUCT-O. | IUCT-O. |
| 3 | IUCT-O. | CHU Hôtel Dieu. | University of Washington. |
| 4 | CHU Hôtel Dieu. | University of Washington. | Hospital Clinic Barcelona. |
| 5 | CHRU-Hôpital Claude Huriez. | Institut Paoli-Calmettes. | Tehran University of MeSc. |
| 6 | CHU Amiens sud. | Hospital Clinic Barcelona. | CHU Hôtel Dieu. |
| 7 | CHU Côte de Nacre. | Tehran University of MeSc. | Zhejiang University. |
| 8 | CHU Dijon. | CHRU-Hôpital Claude Huriez. | F.H. Cancer Research Center. |
| 9 | CHU Haut Lévêque. | CHU Amiens sud. | AMPATH Oncology Institute. |
| 10 | CHU La Mileterie. | CHU Côte de Nacre. | CHRU-Hôpital Claude Huriez. |
| 11 | Clinics universities Saint-Luc. | CHU Dijon. | CHU Amiens sud. |
| 12 | Hematology Hôpital Avicenne. | CHU Haut Lévêque. | CHU Côte de Nacre. |
| 13 | Hospices Civils de Lyon. | CHU La Mileterie. | CHU Dijon. |
| 14 | Université de Toulouse. | Clinics universities Saint-Luc. | CHU Haut Lévêque. |
| 15 | Ankara University. | Hematology Hôpital Avicenne. | CHU La Mileterie. |

From Table4, Table5 and Table6, We note that the smaller tuning parameter $\alpha$ allows the degree centrality to be determined mainly by the number of degrees, while institutions with larger degrees are those in HICs, and as the value of the tuning parameter $\alpha$ increases, some institutions with higher numbers of authors move up in the rankings. In addition to this, we find that for the three different tuning parameters, there are institutions ranked in the top 15 in each case, such as CHU de Toulouse, IUCT-O (short for Institut Universitaire du Cancer de Toulouse-Oncopole), CHU Hôtel Dieu, all of which are French institutions.

These institutions in Figure9 form a non-open cluster, i.e. a large number of French institutions have a lot of mutual cooperation with a few Belgian institutions (which explains their high degrees), but they do not cooperate with institutions from other countries, let alone those from LMICs, as can also be seen from the LMHC column in the Table4. The ranking of the institutions in that cluster above is inflated, considering that cooperation between institutions in HICs and those in LMICs is rare but valuable and that such institutions are more useful for potential future cross-income-level cooperation. So the original degree centrality is not a good choice here.

Table 4: Main metrics of the top 15 institutions at $\alpha$=0.2

| Institution | Country | Type | No.au | Deg | HCHC | LMHC | LMLM | Centrality |
|---|---|---|---|---|---|---|---|---|
| Institut Paoli-Calmettes. | France. | HC | 3 | 70 | 67 | 3 | 0 | 77.44084 |
| CHU de Toulouse. | France. | HC | 5 | 49 | 49 | 0 | 0 | 71.53746 |
| IUCT-O. | France. | HC | 5 | 49 | 49 | 0 | 0 | 71.53746 |
| CHU Hôtel Dieu. | France. | HC | 3 | 49 | 49 | 0 | 0 | 64.7873 |
| CHRU-Hôpital Claude Huriez. | France. | HC | 2 | 49 | 49 | 0 | 0 | 59.83113 |
| CHU Amiens sud. | France. | HC | 2 | 49 | 49 | 0 | 0 | 59.83113 |
| CHU Côte de Nacre. | France. | HC | 2 | 49 | 49 | 0 | 0 | 59.83113 |
| CHU Dijon. | France. | HC | 2 | 49 | 49 | 0 | 0 | 59.83113 |
| CHU Haut Lévêque. | France. | HC | 2 | 49 | 49 | 0 | 0 | 59.83113 |
| CHU La Mileterie. | France. | HC | 2 | 49 | 49 | 0 | 0 | 59.83113 |
| Clinic university Saint-Luc. | Belgium. | HC | 2 | 49 | 49 | 0 | 0 | 59.83113 |
| Hematology Hôpital Avicenne. | France. | HC | 2 | 49 | 49 | 0 | 0 | 59.83113 |
| Hospices Civils de Lyon. | France. | HC | 2 | 49 | 49 | 0 | 0 | 59.83113 |
| Université de Toulouse. | France. | HC | 2 | 49 | 49 | 0 | 0 | 59.83113 |
| Ankara University. | Turkey. | LMIC | 2 | 50 | 0 | 42 | 8 | 51.14623 |

Table 5: Main metrics of the top 15 institutions at $\alpha$=0.5

| Institution | Country | Type | No.au | Deg | HCHC | LMHC | LMLM | Centrality |
|---|---|---|---|---|---|---|---|---|
| CHU de Toulouse. | France. | HC | 5 | 49 | 49 | 0 | 0 | 126.1943 |
| IUCT-O. | France. | HC | 5 | 49 | 49 | 0 | 0 | 126.1943 |
| CHU Hôtel Dieu. | France. | HC | 3 | 49 | 49 | 0 | 0 | 98.49873 |
| University of Washington. | USA. | HC | 19 | 27 | 13 | 14 | 0 | 96.79359 |
| Institut Paoli-Calmettes. | France. | HC | 3 | 70 | 67 | 3 | 0 | 90.11104 |
| Hospital Clinic Barcelona. | Spain. | HC | 25 | 23 | 20 | 3 | 0 | 83.20457 |
| Tehran University of MeSc. | Iran. | LMIC | 19 | 26 | 0 | 10 | 16 | 82.37718 |
| CHRU-Hôpital Claude Huriez. | France. | HC | 2 | 49 | 49 | 0 | 0 | 80.72794 |
| CHU Amiens sud. | France. | HC | 2 | 49 | 49 | 0 | 0 | 80.72794 |
| CHU Côte de Nacre. | France. | HC | 2 | 49 | 49 | 0 | 0 | 80.72794 |
| CHU Dijon. | France. | HC | 2 | 49 | 49 | 0 | 0 | 80.72794 |
| CHU Haut Lévêque. | France. | HC | 2 | 49 | 49 | 0 | 0 | 80.72794 |
| CHU La Mileterie. | France. | HC | 2 | 49 | 49 | 0 | 0 | 80.72794 |
| Clinic university Saint-Luc. | Belgium. | HC | 2 | 49 | 49 | 0 | 0 | 80.72794 |
| Hematology Hôpital Avicenne. | France. | HC | 2 | 49 | 49 | 0 | 0 | 80.72794 |

Table 6: Main metrics of the top 15 institutions at $\alpha$=0.8

| Institution | Country | Type | No.au | Deg | HCHC | LMHC | LMLM | Centrality |
|---|---|---|---|---|---|---|---|---|
| CHU de Toulouse. | France. | HC | 5 | 49 | 49 | 0 | 0 | 222.6106 |
| IUCT-O. | France. | HC | 5 | 49 | 49 | 0 | 0 | 222.6106 |
| University of Washington. | USA. | HC | 19 | 27 | 13 | 14 | 0 | 208.2267 |
| Hospital Clinic Barcelona. | Spain. | HC | 25 | 23 | 20 | 3 | 0 | 179.9694 |
| Tehran University of MeSc. | Iran. | LMIC | 19 | 26 | 0 | 10 | 16 | 164.5535 |
| CHU Hôtel Dieu. | France. | HC | 3 | 49 | 49 | 0 | 0 | 149.7516 |
| Zhejiang University. | China. | LMIC | 14 | 5 | 0 | 0 | 5 | 132.6317 |
| F.H. Cancer Research Center. | USA. | HC | 20 | 5 | 4 | 1 | 0 | 121.5864 |
| AMPATH Oncology Institute. | Kenya. | LMIC | 18 | 13 | 0 | 1 | 12 | 116.6997 |
| CHRU-Hôpital Claude Huriez. | France. | HC | 2 | 49 | 49 | 0 | 0 | 108.9232 |
| CHU Amiens sud. | France. | HC | 2 | 49 | 49 | 0 | 0 | 108.9232 |
| CHU Côte de Nacre. | France. | HC | 2 | 49 | 49 | 0 | 0 | 108.9232 |
| CHU Dijon. | France. | HC | 2 | 49 | 49 | 0 | 0 | 108.9232 |
| CHU Haut Lévêque. | France. | HC | 2 | 49 | 49 | 0 | 0 | 108.9232 |
| CHU La Mileterie. | France. | HC | 2 | 49 | 49 | 0 | 0 | 108.9232 |

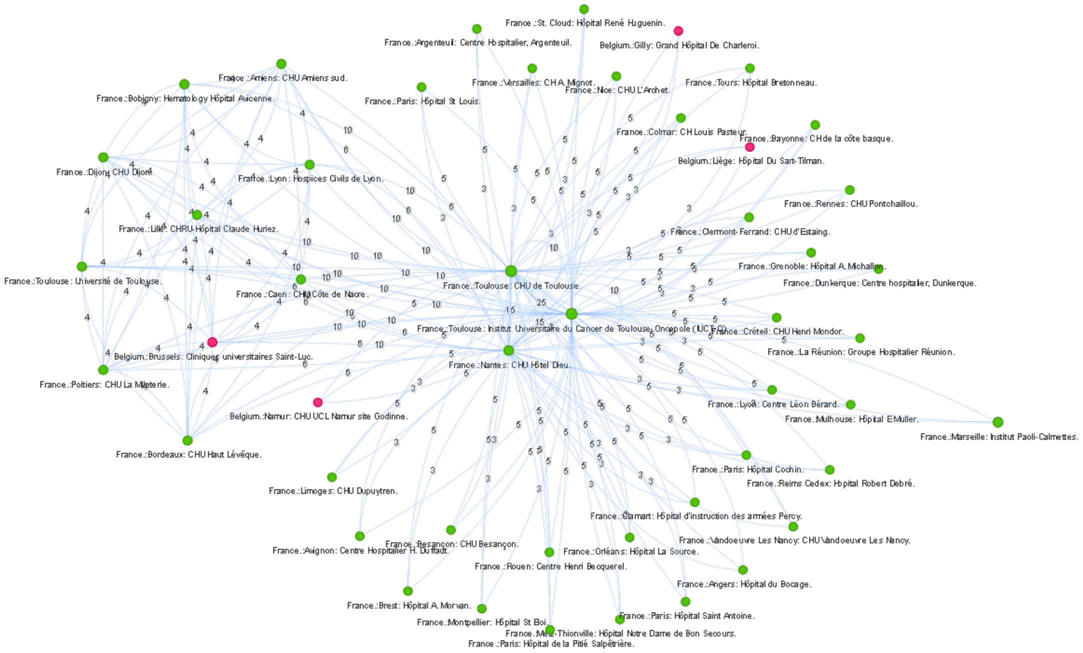


Figure 9:
**Sub-graph of the French cluster.** Edges with weights less than or equal to 2 have been omitted from the network to focus on highly connected institutions.
